# Serum responses of children with Kawasaki Disease against SARS-CoV-2 proteins

**DOI:** 10.1101/2020.05.24.20111732

**Authors:** Arthur J Chang, Michael Croix, Patrick Kenney, Sarah Baron, Mark D Hicar

## Abstract

Recently, numerous reports have suggested association of pediatric Coronavirus Disease 2019 (COVID-19) cases and Kawasaki Disease (KD). KD is a major cause of childhood acquired heart disease and vasculitis in the pediatric population. Epidemiological patterns suggest KD is related to an infectious agent; however, the etiology remains unknown1. As past reports have considered other coronaviruses to be related to KD2,3, these reports of pediatric COVID-19 related inflammatory disorder cases leads to the hypothesis of potential cross-coronavirus reactivity that would account for the past controversial proposals of other coronaviruses and these new cases. We sought to address this hypothesis by assessing the antigen targeting of biobanked plasma samples of febrile children, including those with KD, against SARS-CoV-2 proteins.

## Introduction

Recently, numerous reports have suggested association of pediatric Coronavirus Disease 2019 (COVID-19) cases and Kawasaki Disease (KD). KD is a major cause of childhood acquired heart disease and vasculitis in the pediatric population. Epidemiological patterns suggest KD is related to an infectious agent; however, the etiology remains unknown^1^. As past reports have considered other coronaviruses to be related to KD^2,3^, these reports of pediatric COVID-19 related inflammatory disorder cases leads to the hypothesis of potential cross-coronavirus reactivity that would account for the past controversial proposals of other coronaviruses and these new cases. We sought to address this hypothesis by assessing the antigen targeting of biobanked plasma samples of febrile children, including those with KD, against SARS-CoV-2 proteins.

## Methods

We collected blood samples from SARS-CoV-2 PCR positive patients as previously described^4^ under the University at Buffalo IRB approved STUDY00004340. The ELISA protocol used followed a recently published SARS-CoV-2 related protoco^5^ with minor adjustments (use of Goat Anti-Human Ig-HRP (Southern Biotech) secondary and TMB Ultra (Thermo Fisher) as developer. This was supported by SUNY Research Seed Grant Program 2019-2020. Statistical analysis was performed using GraphPad Prism 8 (v8.4.2). KD subject samples were collected under approval of the University at Buffalo (UB) IRB (MODCR00000185) with funding support by the Wildermuth Foundation through the Variety Club of Buffalo.

## Results

Three ELISAs were developed using commercially available proteins (Sino Biologics SARS-CoV-2 proteins: receptor binding domain (RBD) Cat#40592-V08H; Nucleocapsid Protein (NP) Cat# 40588-V08B; and Spike Protein, S1+S2 ECD, Cat#40589-V08B1). These showed high specificity of binding when tested with rabbit polyclonal antibodies (also obtained from Sino Biologics, data not shown). We then tested our 14 SARS-CoV-2 PCR positive patients’ plasma samples (**Figure 1**: dilution of 1:50 is shown). Enrollees with blood samples taken after day 10 of illness were positive for all three antigens. There was variability in positivity of the early samples (day 3-10 days of illness).

**Figure 1:**
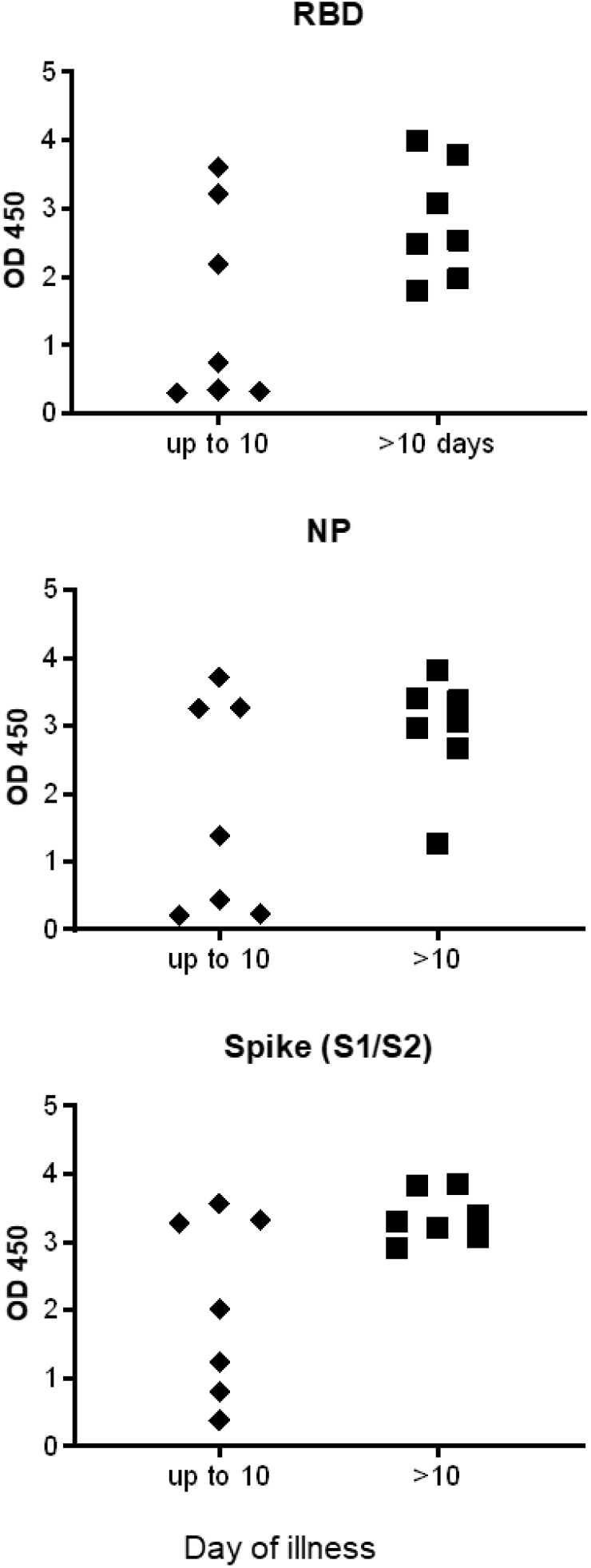
SARS-CoV-2 protein recognition by COVID-19 patients. 14 patients with PCR confirmed COVID-19 had plasma diluted 1:50. Raw OD data from ELISAs are presented.

We then tested plasma samples, collected from 2013-2019, of 36 febrile controls, 24 pre-intravenous immunoglobulin (IVIG) KD samples, 12 post-IVIG KD samples, and 3 convalescent KD samples (**Figure 2**). A SARS-CoV-2 PCR positive patient’s convalescent serum was assayed on each plate as an internal control, and levels at 1:50 and 1:500 are shown as high and low positive ranges on the figures. Targeting of the RBD was most specific as a number of positive reactions to NP and Spike were observed in the febrile controls. We did not see any significant reactivity to SARS-CoV-2 RBD, NP, or Spike protein from in the KD samples. In those KD samples, IVIG infusion did seem to associate with higher binding on all three assays. The ratios of the means of post-IVIG to pre-IVIG were 1.47, 1.73, and 1.75 respectively for RBD, NP, and Spike proteins.

**Figure 2:**
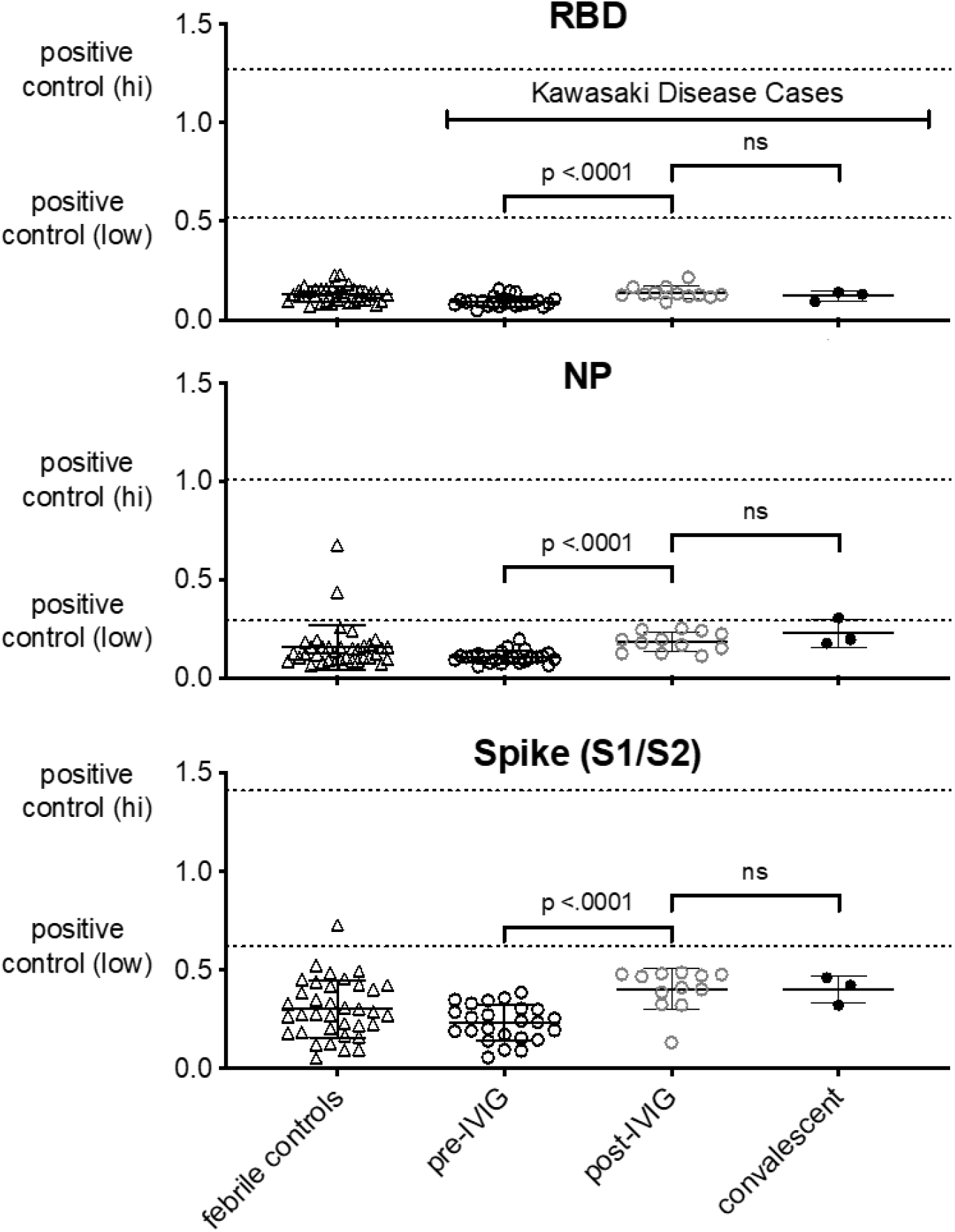
Samples from Kawasaki Disease lack cross reactivity to SARS-CoV-2. Raw OD data presented, with plasma diluted 1:50. Internal positive control depicted at dilution at 1:50 (positive control (hi)) and 1:500 (positive control (low)).

## Discussion

From this study, we would conclude that there is not a non-specific global cross-reactivity from previous coronaviruses contributing to these pediatric COVID-19 related inflammatory disorder cases.

Changes in binding observed post-IVIG infusion imply non-specific interaction due to higher level of circulating immunoglobulins after IVIG infusion. While this change was statistically significant in all 3 assays, NP and Spike were more pronounced. This suggests some cross reactivity in IVIG preparations to other coronaviruses for those antigens. As a majority of our post-IVIG samples were collected at day 10, we would have expected more significant elevation (as highlighted in **Figure 1**) if this post-IVIG increase was from seroconversion.

We also confirm other observations about positive serology >10 days from symptom onset as compared to subjects who were tested ≤10 days from symptom onset^6^. Additional studies in children are needed to see if this pattern applies to children as well.

As both COVID-19 and KD appear to incite a large inflammatory response in certain children, continued research is needed to provide the pathophysiologic explanation of these events in SARS-CoV-2 and Kawasaki disease.

## Data Availability

Data available by contacting author.

## Clinical Trial Registration (if any)

none

**Abbreviations**
Kawasaki Disease: (KD)
Coronavirus Disease 2019: (COVID-19)
intravenous immunoglobulin: (IVIG)

## Contributors statement

Dr. Chang drafted the initial manuscript, and reviewed and revised the manuscript.

Drs. Croix and Kenney collected data, and reviewed and revised the manuscript.

Sarah Baron designed the study, collected data, and reviewed and revised the manuscript.

Dr. Hicar conceptualized and designed the study, coordinated and supervised data collection, and critically reviewed the manuscript for important intellectual content.

All authors approved the final manuscript as submitted and agree to be accountable for all aspects of the work.

